# Detection of monkeypox viral DNA in a routine wastewater monitoring program

**DOI:** 10.1101/2022.07.25.22278043

**Authors:** Marlene K. Wolfe, Dorothea Duong, Bridgette Hughes, Vikram Chan-Herur, Bradley J. White, Alexandria B. Boehm

## Abstract

Wastewater represents a composite biological sample from the entire contributing population. People infected with monkeypox excrete monkeypox virus DNA via skin lesions, saliva, feces and urine and these can enter the wastewater via toilets, sinks, and shower drains. To test whether monkeypox can be detected and monitored in wastewater during a period when publicly reported monkey cases in the region were increasing, we deployed digital PCR assays that target genomic DNA from the monkeypox virus in our routine, ongoing wastewater surveillance program in the Greater Bay Area of California, USA. We measured monkeypox virus DNA daily in settled solids samples from nine wastewater plants over the period of approximately 4 weeks. During that period, we detected monkeypox virus DNA in wastewater solids at nearly all the wastewater plants we routinely sample. Frequency of occurrence and concentrations were highest at plants serving San Francisco County. To confirm the presence of monkeypox DNA, we used two assays that target distinct sequences on the monkeypox genome on a subset of samples and results from both assays were in close agreement strongly suggesting true positives in the wastewater. Additionally, we show that concentrations of monkeypox DNA is 10^3^ times higher in the solid fraction compared to the liquid fraction of wastewater on a mass-equivalent basis.

## Introduction

Monkeypox virus (MPXV), of the Orthopoxvirus genus, is endemic in Western and Central Africa where infection has been primarily linked to zoonotic transmission from small mammals. Sporadic cases and outbreaks linked to travel to or animals imported from endemic regions have been recognized in non-endemic countries since the first identification of the disease in 1970. In early May 2022, cases of monkeypox infection without association to Western or Central Africa were reported in multiple European countries^1^. Within weeks, cases had been identified in the United States and a dozen countries around the world. By late July, the WHO declared a public health emergency of international concern. Unlike historical outbreaks, the current global outbreak features human-to-human transmission ^2,3^, and cases with no association with each other have been identified, suggesting unrecognized community transmission.

Since the first cases were reported, there has been a rapid scale-up of public health infrastructure to detect circulating disease in an effort to mitigate spread, including substantial increases in testing and efforts to educate clinicians and the public^4^. However, practical access to and utilization of testing is limited by availability of appropriate laboratory safety equipment and vaccinations for laboratory staff, clinical awareness of a non-specific disease that is novel for most clinicians, stigma of a disease that has mostly been reported in gay and bisexual men, and the potential for asymptomatic cases. As such, alternative public health surveillance approaches not reliant on individual polymerase chain reaction (PCR) testing, such as through wastewater surveillance, provide an attractive means to understand emerging epidemiology of monkeypox transmission in local communities and provide situational awareness for public health and clinicians.

The use of wastewater as a public health surveillance tool to monitor trends in infectious diseases has grown rapidly because wastewater acts as a composite biological sample capturing inputs from anyone who is connected to a sewer network. Many infections result in shedding in ways that reliably end up in wastewater, including in urine and feces but also through oral and nasal secretions and sloughing of skin. Wastewater has also been long established as a public health tool to monitor for poliovirus circulation, and monitoring of SARS-CoV-2, the causative virus of Coronavirus disease 2019 (COVID-19), in wastewater has been reliably used by public health throughout the COVID-19 pandemic. Concentrations of SARS-CoV-2 viral RNA are strongly correlated with case incidence ^5–7^, and recent studies show this is also the case for respiratory viruses such as Influenza A and RSV ^8,9^. Evidence for these use cases have been established over the years, improving interpretation of results and increasing confidence for use in public health.

Rapidly leveraging wastewater surveillance infrastructure may enhance surveillance for an emerging infectious disease such as monkeypox in real-time with minimal changes in sample collection procedures, however it is unclear how effectively these systems can be used for new outbreaks where viral shedding profiles are still being delineated. Observational studies of patients infected during the current monkeypox outbreak (summer 2022) confirms shedding of viral DNA from saliva, feces, urine, semen, and skin lesions^10,11^. Authors did not report concentrations or mass of viral DNA in their specimens, however reported Cq values from qPCR measurements were low, suggesting high concentrations of MPXV DNA even on day 16 after symptom onset^10^. These data are consistent with reports of MPXV shedding from previous outbreaks in humans^12^ and experimental studies in animals^13–16^ that consistently show shedding via these excretions. Related viruses including smallpox^17^ have been shown to be excreted in urine. Together, this evidence suggests that MPXV DNA is likely to appear in wastewater. Based on a systematic review of the literature^18^, no study to date has documented the persistence of orthopoxviruses in wastewater; however data on Vaccinia virus in raw freshwater and marine waters indicate that it can persist for days^18^, suggesting orthopoxviruses may persist in wastewater as it transits from homes to nearby wastewater treatment plants.

The goal of this work was to investigate whether an assay could be rapidly adapted and deployed for surveillance of monkeypox in multiple sewersheds in California, and to investigate whether MPXV DNA can be consistently detected in municipal wastewater.

## Methods

### Molecular Assay choice for monkeypox

To detect MPXV DNA in wastewater, we used assays developed by the United States Centers for Disease Control and Prevention (CDC)^19^. The G2R_G assay, which targets a a region of the OPG002 gene common to all MPXV sequences, was used on all samples. A subset of samples (described further below) were also assayed for MPXV DNA using the G2R_WA assay which targets a separate region of the OPG002 gene that is specific to the West Africa clade. The choice of these assays was confirmed by alignment with sequences from the 2022 outbreak.

### Wastewater samples for monkeypox surveillance

Each day between 19 June 2022 and 20 July 2022, settled solids were collected in sterile containers from nine publicly owned treatment works (POTWs) in the Greater San Francisco Bay Area of California including two POTWs in the Sacramento Area (Table S1). The POTWs treat wastewater from between 66,622 and 1,480,00 people; further details of eight of the POTWs, as well as specific sample collection processes are provided in Wolfe et al.^5^. The ninth POTW, SEP, is located in San Francisco county and treats sewage from 650,000 people. Solids samples were couriered to a laboratory on the same day they were collected and processed immediately, unless otherwise specified (Table S2, with results available within 24 hour of sample collection. A total of 287 solids samples were collected for analysis in this study.

After receipt at the laboratory, nucleic acids were extracted and purified from the solids using previously published methods^5,20–22^. In brief, dewatered solids were suspended in a buffer and homogenized, and then 10 replicate aliquots of the buffer were subjected to nucleic acid extraction and purification, followed by inhibitor removal using commercial kits (Chemagic Viral DNA/RNA 300 Kit H96 for Chemagic 360 (PerkinElmer, Waltham, MA) and Zymo OneStep-96 PCR Inhibitor Removal kit (Zymo Research, Irvine, CA)). Nucleic acids from each of the 10 replicates were used undiluted as template in a single 20 μL digital droplet PCR well (10 replicate wells total per sample); results from the wells were merged to determine the concentration of the G2R_G target. PCR cycling conditions and further details of ddPCR are provided in the SI. We also measured PMMoV RNA gene concentrations; as well as recovery of spiked-in bovine coronavirus RNA following methods outlined in detail elsewhere and available on protocols.io. Additionally, negative and positive extraction controls, and negative and positive PCR controls were included in each plate. Gene blocks were used as controls for the G2R_G assay, other controls are described elsewhere^5^. Nucleic-acids were not stored prior to analysis with the exception of some nucleic-acids from SEP which were stored at -80°C less than two weeks and thus subjected to a freeze thaw prior to G2R_G analysis (Table S2).

A subset of samples from two POTWs (OSP and SEP, Table S2) with the highest rates of G2RR2_G detection were processed for the second MP genomic target, G2RR2_WA, using the same replication and QA/QC as described for G2RR2_G. A gene block was used as the G2RR2G_WA positive control. PCR cycling conditions are provided in the SI.

### Liquid influent samples

Samples of 24-h composited influent were collected in sterile containers on ∼7 consecutive days (Table S2) from two POTWs (OSP and SEP) that had the highest rates of detection of the G2R_G target in order to compare the concentrations obtained using liquid wastewater to those obtained using settled solids. Samples were couriered to the laboratory on the same day they were collected and stored at 4°C for between 1 and 7 d before being processed together in one batch. For each sample, 10 replicate aliquots were processed using an affinity-based capture method with magnetic hydrogel Nanotrap Particles with Enhancement Reagent 1 (Ceres Nanosciences, Manassas, VA) on 10mL of sample to concentrate viral particles using a KingFisher Flex system. RNA was then extracted from the each concentrated aliquot using the MagMAX Viral/Pathogen Nucleic Acid Isolation Kit (Applied Biosystems, Waltham, MA) on the KingFisher Flex platform to obtain purified nucleic acids which were then process through a Zymo OneStep-96 PCR Inhibitor Removal kit (Zymo Research, Irvine, CA)). Nucleic acids were used undiluted as template in digital droplet PCR to measure concentrations of G2R_G and G2R_WA targets, as well as PMMoV and BCoV recovery following the same protocol as described for the solids.

### Statistical analysis

We assessed the relationship between MPXV DNA concentrations in measurements taken from liquid and solids samples and the relationship between results from the G2R_G and G2R_WA assays on the same sample using Kendall’s tau. Non-parametric methods were chosen as data were not normally distributed (Shapiro-Wilk test). A paired Wilcoxon Sign Rank test was then used to assess the difference in results between liquids and solids, and between the G2R_G and G2R_WA assays used on the same samples. Measurements below the detection limit were replaced with a value approximately half the detection limit (500 cp/g) for analysis. All statistical analysis was performed in RStudio (version 2021.09.2).

## Results and Discussion

Wastewater monitoring programs can function as a flexible platform that can be rapidly adapted to track new targets, including emerging pathogens. We were able to implement monkeypox testing across sites that were part of a monitoring network providing daily samples, so samples that are routinely tested for SARS-CoV-2, Influenza A, and RSV RNA (among other targets) could also be utilized for monkeypox surveillance. We found that MPXV DNA was consistently detected in wastewater samples across the majority of sites (8/9) monitored during the study period from 6/19/22 - 7/20/22 (dates in format month/day/year) with increasing rates of detection of MPXV DNA concentrations (Table 1, Fig 1). Concentrations of MPXV DNA (the G2R_G target) ranged from non-detect to 24,114 copies/g dry weight of wastewater solids (on 7/19/22 at SEP). Positive and negative controls were all positive and negative respectively, BcoV recoveries were higher than 10%, and PMMoV concentrations were within an expected range for each POTW. This indicates assays performed efficiently and without contamination.

**Fig 1.**
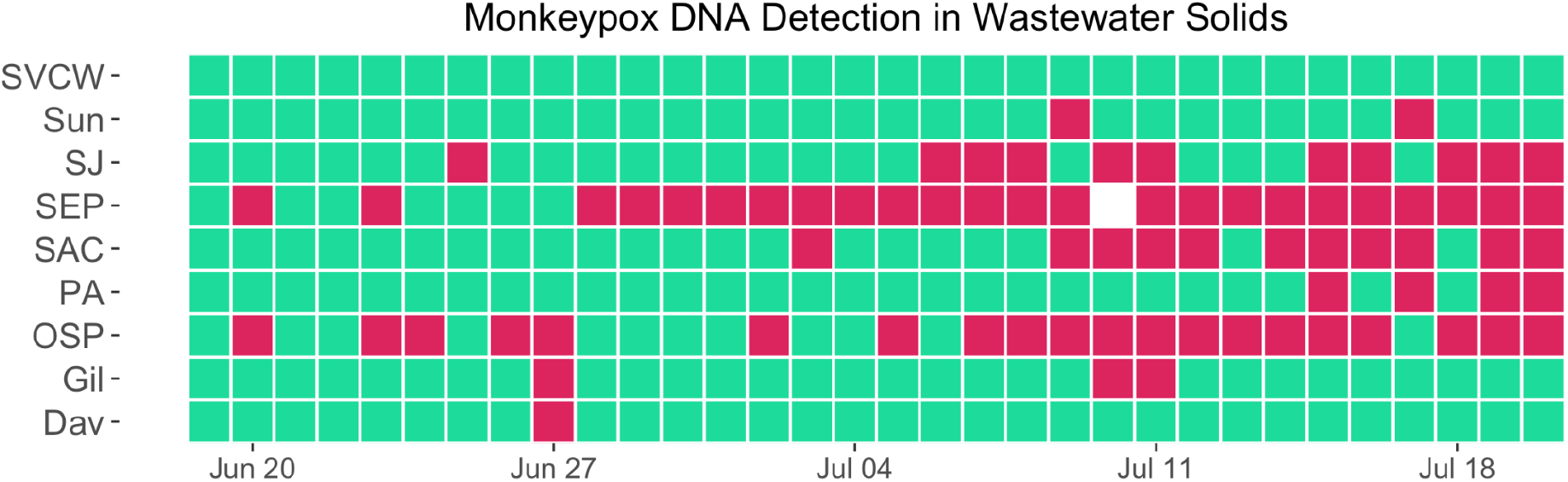
Heatmap of positive and negative results from all POTWs. Plant abbreviations are defined in Table S1. Red indicates a positive sample, green indicates a negative sample. White indicates no sample collected (on 7/10/22 at SEP a sample was missed by the operators).

The first samples to test positive were two samples from 6/20/22 taken from each of the facilities serving the city of San Francisco (OSP and SEP). Daily samples from these sites were sporadically positive for the following 1-2 weeks, after which samples were consistently positive for MPXV DNA and concentrations of MPXV DNA increased (Fig. 2). The next positive sites were SJ (sample from 6/25/22) and SAC (sample from 7/3/22). These sites also followed a pattern of sporadic positives followed by increasing rates of positive samples and concentrations. Samples from SUN, PA, GIL, and DAV also tested positive, but did not have increasing rates or trends as of the writing of this manuscript.

**Fig 2.**
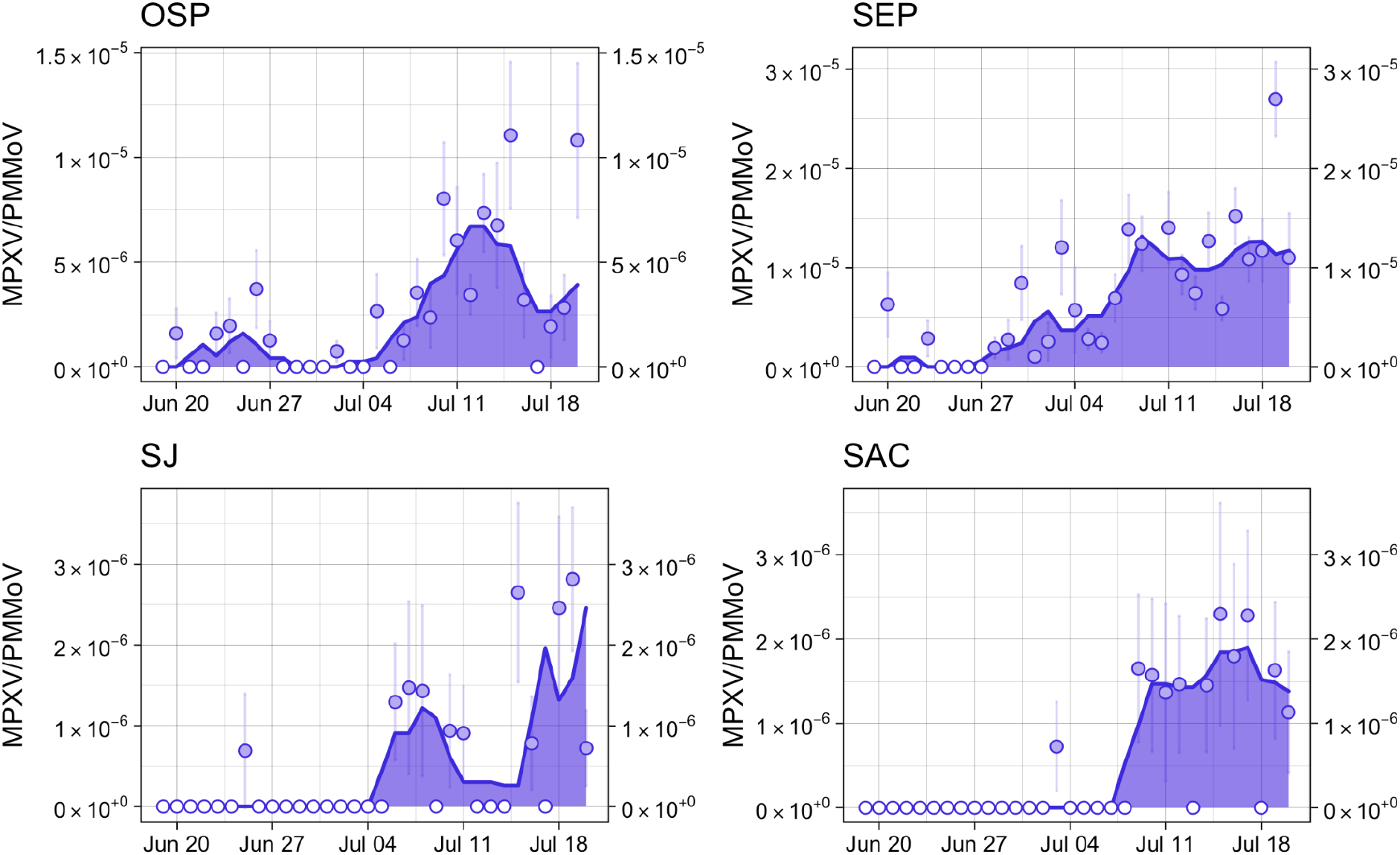
Time series of wastewater concentrations (concentration of MPXV DNA normalized by concentration of PMMoV RNA) at select POTWs with >3 positive detections during the study time period. The area under the curve represents the 5 day trimmed average of MPXV DNA cp/g over PMMoV cp/g in wastewater. Points represent daily values; open circles indicate non-detects. Error bars represent standard deviations and include Poisson error and variability among the 10 replicates (68% confidence intervals reported by the instrument software as “total error”).

### G2R_WA Assay

To provide a second line of evidence for MPXV DNA detection, a subset of samples were run with the second monkeypox specific assay, the G2R_WA assay specific to a target in the West African clade. Results from the G2R_G assay and the G2R_WA assay were significantly associated (Kendall’s tau = 0.88, n = 34, p < 0.001) and there was no significant difference between the two measurements (Wilcoxon signed rank test, n = 34, p = 0.16) (Figure 3). Consistent results across the two assays provides confidence that MPXV DNA detections using the G2R_G assay are true positives.

**Fig 3.**
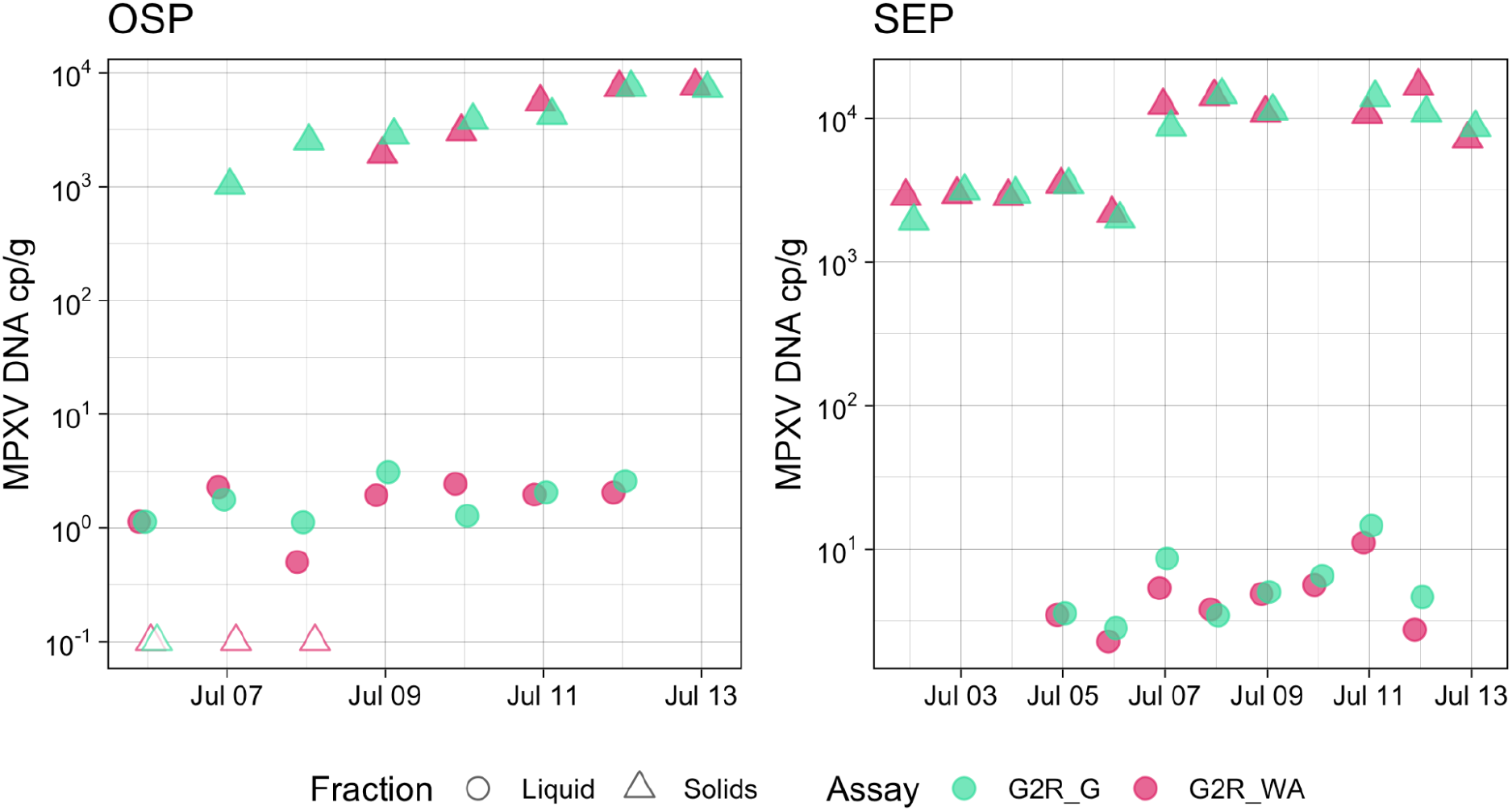
Concentrations of G2R_G and G2R_WA in wastewater solids and liquid influent at OSP and SEP POTWs. Units are per gram for each (one ml of wastewater influent ∼ 1 g of wastewater influent).

### Solids and Liquids Comparison

Our methods focus on the use of wastewater solids, as previous research indicates that many viruses are highly concentrated in solids in wastewater. We compared concentrations of MPXV DNA in paired liquid and solids samples from OSP and SEP over a period of one week. Of the liquid samples, 15/15 were positive for the G2R_G assay and 15/15 were positive for the G2R_WA assay. Concentrations in all samples were near the detection limit. There was a significant association between the results from liquids and solids (Kendall’s tau = 0.52, n = 28, p = 0.00012). Concentrations of MPXV DNA in solids by either assay were significantly higher than those in liquids on a per mass basis (Wilcoxon signed rank test, n = 28, p<0.001), with about a 10^3^ higher concentration of viral DNA observed per g or mL in solids.

Historically mostly limited to use for monitoring outbreak of polio and similar enteric diseases, wastewater monitoring has expanded greatly in popularity and gained widespread use during the COVID-19 pandemic. Robust surveillance for monkeypox can not only enable the targeting of public health resources towards communities experiencing outbreaks, it can help raise awareness among health care professionals to prepare them to recognize and manage cases of monkeypox in their practice.

## Data Availability

All data produced in the present study are available at wbe.stanford.edu and upon request to the authors.

https://wbe.stanford.edu

## Acknowledgements

This work is supported by a gift from the CDC Foundation. Numerous people contributed to sample collection, including Srividhya Ramamoorthy (Sac), Michael Cook (Sac), Ursula Bigler (Sac), James Noss (Sac), Lisa C. Thompson (Sac), Payal Sarkar (SJ), Ryan Batjiaka (Ocean and SEP), Melanie Wong (SEP), Lily Chan (Ocean), the Oceanside and Southeast plant operations personnel, Karin North (PA), Armando Guizar (PA), Saeid Vaziry (Gil), Chris Vasquez (Gil), Alo Kauravlla (Sun), Maria Gawat (SVCW), Tiffany Ishaya (SVCW), Eric Hansen (SVCW), and Jeromy Miller (Dav). We acknowledge Alexander Yu for his partnership in this work. This study was performed on the ancestral and unceded lands of the Muwekma Ohlone people. We pay our respects to them and their Elders, past and present, and are grateful for the opportunity to live and work here. Dorothea Duong, Bridgette Hughes, Vikram Chan-Herur, and Bradley White are employees of Verily Life Sciences.

