## Supplementary Information for "Detection of monkeypox viral DNA in a routine wastewater monitoring program"

**G2RR2_G and G2RR2_WA ddPCR detailed methods.** dd-PCR was performed on 20 µl samples from a 22 µl reaction volume, prepared using 5.5 µl template, mixed with 5.5 µl of One-Step RT-ddPCR Advanced Kit for Probes (Bio-Rad 1863021), 2.2 µl of 200 U/µl Reverse Transcriptase, 1.1 µl of 300 mM DTT and primers and probes at a final concentration of 900 nM and 250 nM respectively. Primer and probes for assays were purchased from Integrated DNA Technologies (IDT, San Diego, CA) (Table S3). We used one step RT-ddPCR mastermix so that we could multiplex the samples with genomic RNA (gRNA) targets of other viruses as part of our regional wastewater monitoring program. The G2R_G assay was multiplexed with an assay targeting gRNA of human metapneumovirus and a mutation in SARS-CoV-2 Omicron BA.5 (HV69-70) (results for other assays not provided herein). The G2R_WA assay was multiplexed with assays targeting gRNA of human rhinovirus and influenza B virus (data from these RNA viruses is not included herein). We assayed the same nucleic-acid extracts from wastewater samples for G2R_G using ddPCR mastermix and one-step RT-ddPCR mastermix in conjunction with a RT step during thermocycling and found results did not differ (Figure SZ).

Droplets were generated using the AutoDG Automated Droplet Generator (Bio-Rad, Hercules, CA). PCR was performed using Mastercycler Pro (Eppendforf, Enfield, CT) with with the following cycling conditions: reverse transcription at 50°C for 60 minutes, enzyme activation at 95°C for 5 minutes, 40 cycles of denaturation at 95°C for 30 seconds and annealing and extension at 59°C for 30 seconds, enzyme deactivation at 98°C for 10 minutes then an indefinite hold at 4°C. The ramp rate for temperature changes were set to 2°C/second and the final hold at 4°C was performed for a minimum of 30 minutes to allow the droplets to stabilize. Droplets were analyzed using the QX200 Droplet Reader (Bio-Rad). A well had to have over 10,000 droplets for inclusion in the analysis. All liquid transfers were performed using the Agilent Bravo (Agilent Technologies, Santa Clara, CA).

Thresholding was done using QuantaSoft™ Analysis Pro Software (Bio-Rad, version 1.0.596). In order for a sample to be recorded as positive, it had to have at least 3 positive droplets. Each wastewater sample was run in 10 replicate wells, and each 96-well PCR plate of wastewater samples included PCR positive controls for each target assayed on the plate in 1 well, and PCR NTCs in two wells. PCR positive controls consisted of gene blocks .

Results from replicate wells were merged for analysis. For the wastewater solid samples, three positive droplets across 10 merged wells corresponds to a concentration between ~500-1000 cp/g; the range in values is a result of the range in the equivalent mass of dry solids added to the wells. For the wastewater influent samples, three positive droplets across 10 merged wells corresponds to a concentration between ~1 cp/ml.

Concentrations of RNA targets were converted to concentrations per dry weight of solids in units of copies/g dry weight or copies / ml for influent using dimensional analysis. The dry weight of the dewatered solids was determined by drying in an oven[1]. The total error is reported as standard deviations and includes the errors associated with the Poisson distribution and the variability among the 10 replicates.

Table S1. Some POTW characteristics. See Wolfe et al.[2] or wbe.stanford.edu for more information.

| Plant names and abbreviations | County locations in California, USA | Population served |
|---|---|---|
| San Jose (SJ) | Santa Clara County | 1,458,017 |
| Palo Alto (PA) | Santa Clara County | 213,968 |
| Gilroy (Gil) | Santa Clara County | 110,338 |
| Sunnyvale (Sun) | Santa Clara County | 169,000 |
| Silicon Valley Clean Water (SVCW) | San Mateo County | 110,338 |
| Oceanside (OSP) | San Francisco County | 250,000 |
| Southeast (SEP) | San Francisco County | 650,000 |
| Sacramento (SAC) | Sacramento County | 1,480,000 |
| Davis (Dav) | Davis County | 66,622 |

Table S2. Details of sample collection and storage. Dates are in month/day/year format

| | |
|---|---|
| Dates influent collected at OSP | 7/6/22-7/12/22 |
| Dates influent collected at SEP | 7/5/22 - 7/12/22 |
| Dates SEP solids analyzed for G2R_WA | 7/2/22 - 7/9/22, 7/11/22 - 7/13/22 |
| Dates OSP solids analyzed for G2R_WA | 7/6/22 - 7/13/22 |
| Dates SEP solids nucleic acids stored at -80°C prior to ddPCR analysis | 6/18/22 - 6/30/22 and 7/2/22 - 7/4/22 |
| Dates SEP solids samples not processed immediately and stored at 4°C for between 1 and 7 days | 7/5/22 - 7/14/22 |

Table S3. Primer and probe sequences from Li et al [3].

| Target | Primer/Probe | Sequence |
|---|---|---|
| G2R_G | Forward | GGAAAATGTAAAGACAACGAATACAG |
| | Reverse | GCTATCACATAATCTGGAAGCGTA |
| | Probe | AAGCCGTAATCTATGTTGTCTATCGTGTCC |
| G2R_WA | Forward | CACACCGTCTCTTCCACAGA |
| | Reverse | GATACAGGTTAATTTCCACATCG |
| | Probe | AACCCGTCGTAACCAGCAATACATTT |

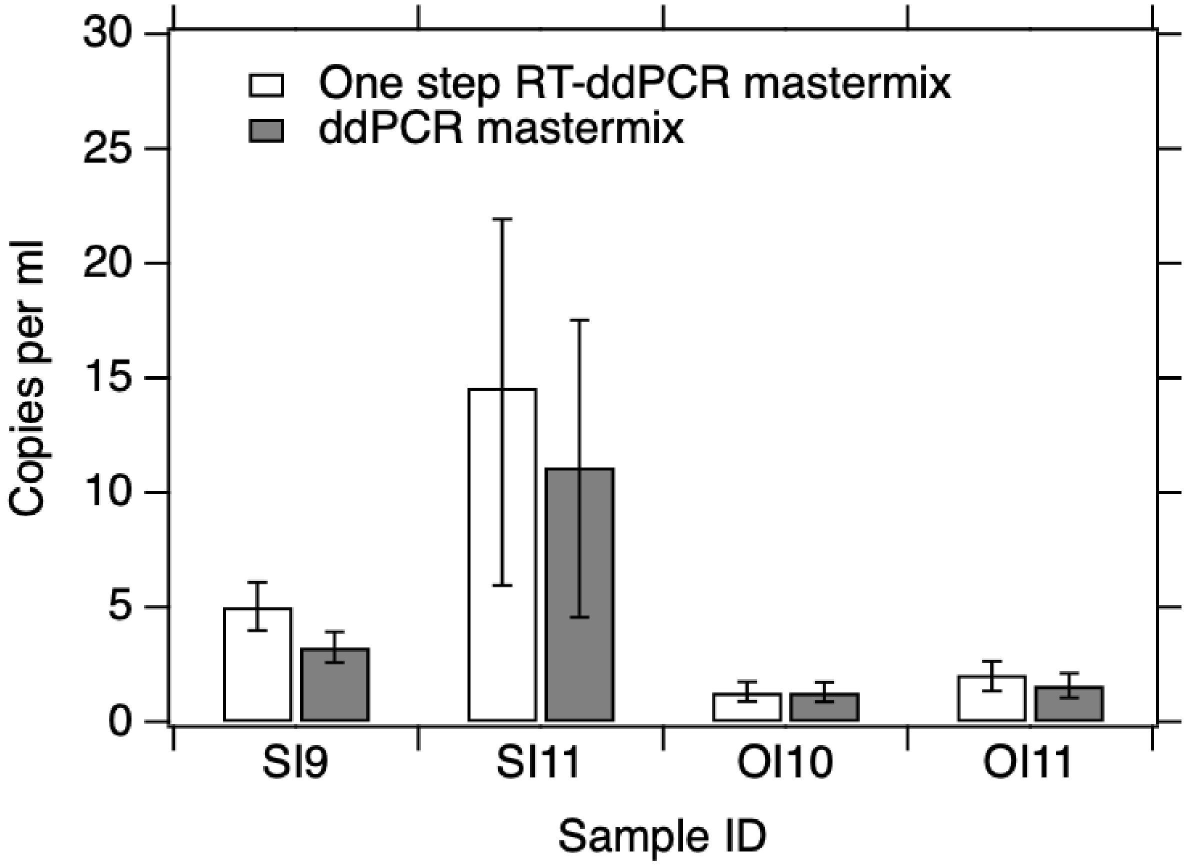


Figure S1. Comparison between wastewater influent nucleic acid extracts run using two different mastermixes using the same methods described in the paper for wastewater sample analysis. The standard deviations represent the total error and includes the Poisson error and the error among replicate wells.


(1) Topol, A.; Wolfe, M.; White, B.; Wigginton, K.; Boehm, A. B. *High Throughput pre-analytical processing of wastewater settled solids for SARS-CoV-2 RNA analyses*. protocols.io. https://www.protocols.io/view/high-throughput-pre-analytical-processing-of-waste-b2kmqcu6 (accessed 2022-07-25).

(2) Wolfe, M. K.; Topol, A.; Knudson, A.; Simpson, A.; White, B.; Vugia, D. J.; Yu, A. T.; Li, L.; Balliet, M.; Stoddard, P.; Han, G. S.; Wigginton, K. R.; Boehm, A. B. High-Frequency, High-Throughput Quantification of SARS-CoV-2 RNA in Wastewater Settled Solids at Eight Publicly Owned Treatment Works in Northern California Shows Strong Association with COVID-19 Incidence. *mSystems 6* (5), e00829-21. https://doi.org/10.1128/mSystems.00829-21.

(3) Li, Y.; Zhao, H.; Wilkins, K.; Hughes, C.; Damon, I. K. Real-Time PCR Assays for the Specific Detection of Monkeypox Virus West African and Congo Basin Strain DNA. *J. Virol. Methods* **2010**, *169* (1), 223–227. https://doi.org/10.1016/j.jviromet.2010.07.012.
